# Cumulative Metabolic Exposure to Hyperglycemia and Risk of Cardiovascular and Limb Events in Peripheral Artery Disease

**DOI:** 10.64898/2026.06.18.26356017

**Authors:** Feroz James, Qiang Li, Medha Somisetty, Cathy Nguyen, Mary Vaughan-Sarrazin, Brian Lund, Shirling Tsai, Kim Smolderen, Richard Hoffman, Shipra Arya, Joshua Beckman, Saket Girotra

## Abstract

**Background:** Although diabetes is a potent risk factor for the development of peripheral artery disease (PAD), the effect of cumulative metabolic exposure to hyperglycemia on risk of cardiovascular or limb events in patients with PAD remains unclear.

**Methods:** The Peripheral Artery Disease: Long-term Survival (PEARLS) is a longitudinal registry of Veterans with newly diagnosed PAD identified using a natural language processing approach. Included patients had ankle brachial index ≤0.9 or toe brachial index ≤0.7, and no history of lower extremity revascularization or major amputation. Among patients with diabetes in this cohort, we assessed cumulative exposure to hyperglycema based on a 24-month rolling average of hemoglobin (Hgb) A1c values, categorized as ≤7%, >7% to ≤8%, and >8%. Multivariable Cox regression models evaluated the association between categories of HgbA1c, modeled as a time-varying exposure, and risk of cardiovascular (CV: myocardial infarction or stroke) and limb (chronic limb threatening ischemia [CLTI] or major amputation) events.

**Results:** Among 45,109 patients with new diagnosis of PAD and pre-existing diabetes, the mean HgbA1c at baseline was 7.5%, with nearly one-third (30.4%) having HgbA1c >8%. The mean age was 70.4 years, 19.8% were Black and 4% were Hispanic. Patients with baseline HgbA1c >8% were younger and compared to those with HgbA1c ≤7%, more likely to have coronary disease, kidney disease, and obesity. Over a median follow up of 4.2 years, 8,306 (18.4%) patients experienced a CV event, and 8,199 (18.2%) experienced a limb event. The adjusted association between HgbA1c and hazard of CV events was 12% higher in patients exposed to HgbA1c >7% to ≤8% (HR 1.12; 95%CI: 1.05-1.18) and 38% higher in those exposed to HgbA1c >8% (HR 1.38; 95%CI: 1.30-1.46), compared to HgbA1c <7%. The association with limb events was even stronger, with 20% and 60% higher hazard in those exposed to >7% to ≤8% (HR 1.20; 95%CI: 1.13-1.28) and HgbA1c >8% (HR 1.60; 95%CI: 1.51-1.70), respectively when compared to HgbA1c ≤7%. These findings were consistent in subgroups based on age and severity of PAD.

**Conclusions:** Among diabetic patients with PAD, cumulatiave metabolic exposure to hyperglycemia is associated with a markedly increased risk of clinical events, especially limb events.

**Clinical Perspective:** *What is new? (maximum 100 words, formatted as 2-3 bullets):*

- Among > 45,000 patients with diabetes and new-onset PAD in a large, multi-center registry, we found a graded positive association between cumulative metabolic exposure to hyperglycemia and risk of cardiovascular (CV) and limb events.
- Poor glycemic control was more strongly associated with limb events than CV events, and this association was consistent in older patients and those with severe PAD.

*What are the clinical implications? (maximum 100 words, formatted as 2-3 bullets).:*

- Patients with PAD have been underrepresented in previous trials evaluating the effect of intensive control of diabetes on CV outcomes.
- Given the remarkably high burden of CV and limb events in those with PAD and diabetes, our findings support the need for a randomized controlled trial to evaluate the role of glycemic control in mitigating vascular disease risk in this population.

## Introduction

Among patients with peripheral artery disease (PAD), diabetes is highly prevalent and associated with a markedly increased risk of cardiovascular (CV) events and progression of PAD to chronic limb threatening ischemia (CLTI) or major amputation.^1,2^ Randomized controlled trials in patients with diabetes have shown that intensive glycemic control reduces the risk of microvascular complications, but evidence demonstrating an improvement in CV events is mixed at best.^3–6^ A modest 10% reduction in CV event risk was observed when the individual trials were combined in a meta-analysis or when follow-up was extended beyond 10 years in the United Kingdom Prospective Diabetes Study (UKPDS).^7,8^

Importantly, patients with PAD were under-represented in the aforementioned clinical trials and none of the trials included limb events as an endpoint. Although observational studies found poor glycemic control to be associated with worse CV and limb outcomes in those with PAD, these studies have been limited to patients undergoing lower extremity revascularization, or have only considered glycemic control via baseline hemoglobin (Hgb) A1c.^9–12^ Yet, adverse metabolic exposure due to hyperglycemia is cumulative and likely varies over time. Few studies have attempted to quantify the the long-term risk associated with cumulative exposure to hyperglycemia on CV and limb events.

To address this gap in knowledge, we leveraged the Peripheral Artery Disease: Long-term Survival (PEARLS) registry of veterans with newly-diagnosed PAD developed using a novel natural language processing (NLP) algorithm. The registry includes rich clinical data including longitudinal HgbA1c values, comprehensive outpatient follow-up and detailed information on CV and limb events. Accordingly, we examined the association between cumulative metabolic exposure to hyperglycemia, modeled using longitudinal HgbA1c values, and incidence of CV and limb events among patients with diabetes and newly-diagnosed PAD.

## Methods

This work was approved by the Institutional Review Board (IRB) and Research & Development Committees at University of Texas Southwestern Medical Center, and the Dallas, Iowa City and Tennessee Valley Health System VA. We followed the Strengthening the Reporting of Observational Studies in Epidemiology (STROBE) checklist while reporting our findings.

### Study Cohort

Details of the PEARLS registry have been previously described, and are also included in the Supplemental Appendix.^13^ Briefly, PEARLS is a longitudinal registry of veterans with a new diagnosis of PAD between 2015-2020. Patients in the registry are identified based on abnormal ankle brachial index (ABI ≤ 0.9) or toe-brachial index (TBI ≤ 0.7), which are extracted from the electronic health record using a validated NLP based algorithm developed by our research team.^14^ In contrast to the low overall diagnostic accuracy of International Classification of Diseases (ICD) codes, the positive predictive value of PAD identification using this algorithm was 92.3%.^14^ Using this approach, we have previously identified 103,748 patients with new diagnosis of PAD during 2015-2020, with index date defined as the date of the first abnormal ABI or TBI result. Data on clinical variables included demographics, co-morbidities, vital signs, laboratory values, and dispensed medications were obtained from relevant domains in the VA Informatics and Computing Infrastructure (VINCI). In addition to data from the Veteran’s Affairs (VA) Corporate Data Warehouse (CDW), clinical endpoints were also identified through linkage of our cohort with VA fee-basis and consolidated datasets (CDS) which include claims data for non-VA services paid by the VA, and all Medicare claims, including fee-for-service inpatient (Part A), outpatient (Part B), managed care (Part C), and pharmacy claims (Part D).^15^ Therefore, these additional data sources ensured a near complete capture of clinical endpoints for patients in our cohort.

We restricted our cohort to those with diabetes, defined as the presence of diabetes as a co-morbidity using the Elixhauser-Charleson comorbidity schema,^16^ filling a prescription for a diabetes medication, or a hemoglobin A1c ≥ 6.5% during the 2 years prior to index date. Participants were excluded if they did not have at least 2 outpatient visits to ensure our cohort was comprised of regular VA users. We also required patients to have at least one HgbA1c value during the 2 years prior to index date, and at least one HgbA1c during the 2 years after index date but before the first event or end of follow-up (Figure S1). The final cohort comprised 45,109 patients with diabetes and newly-diagnosed PAD.

### Study Endpoints and Variables

The study endpoints included a) cardiovascular events, defined as hospitalization for myocardial infarction or stroke, and b) limb events, defined as chronic limb threatening ischemia (CLTI) or major amputation (above ankle). Lower extremity revascularization was not included as an event in the primary analysis because the decision to perform revascularization is often discretionary and does not always imply disease progression. Study endpoints were identified using ICD-10 and CPT codes associated with clinical encounters in the VA, CDS and Medicare data (Table S1). Patients were followed from diagnosis of PAD until first clinical event, death, or the end of 5 years, whichever was earlier.

The primary exposure was cumulative metabolic exposure to hyperglycemia based on a 24-month rolling average of HgbA1c values, categorized as ≤7%, >7% to ≤8% and >8%. Thus, at index (date of PAD diagnosis), all HgbA1c values during the preceding 24 months were averaged and this variable was updated each month of follow-up with the addition of new HgbA1c results when they became available. HgbA1c values falling outside this 24-month period were correspondingly dropped as the rolling average was calculated each month. Study covariates included age, sex, race, ethnicity, smoking, duration of diabetes (in years) and co-morbidities (defined by the Elixhauser-Charlson algorithms) which included hypertension, coronary artery disease, chronic kidney disease, heart failure, cerebrovascular disease, COPD, anemia, valvular heart disease, dementia, chronic liver disease, obesity, cancer, weight loss, arrhythmia, and depression.^17^ We also included PAD severity, defined as mild: 0.8 ≤ ABI ≤ 0.9 or 0.6 ≤ TBI ≤ 0.7; moderate: 0.5 ≤ ABI < 0.8 or 0.4 ≤ TBI < 0.6; severe: 0 ≤ ABI < 0.5 or 0 ≤ TBI < 0.4 in the worst limb. Additional covariates included systolic blood pressure, body mass index (BMI) and low density lipoprotein-cholesterol (LDL-C) values during the 12-month period before the index date from VINCI. Finally, we also included active use of diabetes (metformin, sulfonylureas, insulin, sodium-glucose transport protein inhibitors, glucagon-like peptide-1 receptor agonists, or other), blood pressure (beta blockers, renin-angiotensin aldosterone system inhibitors, diuretics, calcium channel blockers, vasodilators, and others) and statin medications at index date. Medication use was assessed usng VA Pharmacy and Medicare Part D data claims and active use was defined as having filled a prescription for that medication within two times the number of days’ supplied by that prescription before index date.^13^ For example, a patient who filled a 90-day prescription for metformin within 180 days prior to index date would be considered an active metformin user.

### Statistical Analysis

We compared baseline characteristics of study participants stratified by categories of hemoglobin A1c during the baseline period, using a chi-square test for categorical variables and linear regression for continuous variables.

To examine the association between glycemic control and clinical events, separate Cox proportional hazards models were developed for CV and limb events. Models were fitted in monthly increments and HgbA1c was modeled as a time-varying exposure using the average value over the lagging 24 month-period. The models adjusted for all variables listed in the *Study Endpoints and Variables* section. Hazard ratios for CV and limb events were estimated with the HgbA1c ≤7% group as the reference category. Patients were censored if they died prior to an endpoint. A sandwich estimator was used to account for correlation between repeated HgbA1c values. The coefficients for each category were exponentiated to obtain a hazard ratio (HR) with 95% confidence intervals (CI). The proportionality of hazards over time was evaluated using an interaction of time with HgbA1c category, which was not significant.

To further quantify the marginal effect of exposure to each HgbA1c category, we also estimated the adjusted incidence of CV and limb events over a 3-year period using the method of recycled predictions, with separate models for each event type.^18^ This method uses the model estimates to predict the adjusted incidence of CV or limb event for each HgbA1c category, assuming that all patients in the cohort were exposed to the same HgbA1c category during follow-up while holding all other variables constant.

We also examined whether the effect of glycemic control on CV and limb outcomes differed by age (< 70 years vs. ≥ 70 years) and severity of PAD (severe vs. non-severe) at diagnosis by including an interaction of these variables with the primary HgbA1c exposure in another Cox model. The adjusted incidence of clinical events was similarly calculated for these subgroups.

In our primary analysis, we conceptualized that cumulative metabolic exposure to hyperglycemia would affect both CV and limb events over the long-term and therefore included a 24-month rolling average HgbA1c that was updated monthly in the time-varying Cox model. In sensitivity analyses, we examined whether our results were sensitive to this assumption by repeating the analysis using monthly (most recent) values of HgbA1c, also as a time-varying exposure, instead of a 24-month average. All analyses were performed in SAS version 9.4 (SAS Institute, Cary, North Carolina).

## Results

Among 45,109 patients with newly-diagnosed PAD and preexisting diabetes, the mean HgbA1c at baseline was 7.5% (SD 1.5%). At diagnosis of PAD, 19,732 (43.7%) patients had a mean HgbA1c ≤7%, 11,685 (25.9%) patients had HgbA1c >7% to ≤8%, and 13,692 (30.4%) patients had HgbA1c >8%, based on their average HgbA1c values over the preceding 24 months. Over a median follow-up of 4.2 years, the median number of HbA1c values per-patient was 11 (IQR: 7-15) in the analysis of both CV and limb events. Among those with baseline HgbA1c <7%, 80.4% remained in the same category at 2 years of follow-up using the 24-month rolling average of HgbA1c (Table S2). However, among those in HgbA1c >7% to 8% and >8%, only 48.2% and 63.5%, respectively remained in the same HgbA1c category at 2 years of follow-up, supporting the use of a time-varying model.

Table 1 shows characteristics of the overall cohort and stratified by baseline HgbA1c. The mean age was 70.4 years, 97.8% were men, 19.8% were Black and 4% were Hispanic. Nearly 20% had severe PAD and more than 60% were either current or former smokers. There was a high prevalence of co-morbidities, especially hypertension (93.0%), coronary artery disease (48.4%), heart failure (25.9%), chronic kidney disease (29.1%) and obesity (34.2%). Patients with HgbA1c >8% during the baseline period were younger, more likely to be Hispanic, and have a higher prevalence of severe PAD, coronary artery disease, heart failure and chronic kidney disease compared to those with HgbA1c ≤ 7%. Differences in other co-morbidities by baseline HgbA1c were statistically significant but small in magnitude.

**Table 1.** Patient Characteristics of Study Cohort Stratified by Baseline Hemoglobin A1c.

|  | Overall | Categories of Baseline Hemoglobin A1c values |  |  |  |
| --- | --- | --- | --- | --- | --- |
|  | N=45,109 | ≤7%<br>N=19,732 | >7% to 8%<br>N=11,685 | >8%<br>N=13,692 | P value |
| **Variable** |  |  |  |  |  |
| Mean Age, years (SD) | 70.4 (8.3) | 71.4 (8.3) | 71.1 (8.0) | 68.4 (8.3) | <0.001 |
| Male Sex | 44,118 (97.8%) | 19,281 (97.7%) | 11,452 (98.0%) | 13,385 (97.8%) | 0.21 |
| Race |  |  |  |  | <0.001 |
| White | 32,874 (72.9%) | 14,254 (72.2%) | 8,878 (76.0%) | 9,742 (71.2%) |  |
| Black | 8,928 (19.8%) | 4,099 (20.8%) | 1,996 (17.1%) | 2,833 (20.7%) |  |
| Other/Unknown | 3,307 (7.3%) | 1,379 (7.0%) | 811 (6.9%) | 1,117 (8.2%) |  |
| Hispanic Ethnicity | 1,797 (4.0%) | 599 (3.0%) | 451 (3.9%) | 747 (5.5%) | <0.001 |
| PAD Severity |  |  |  |  | <0.001 |
| Mild | 14,529 (32.2%) | 6,586 (33.4%) | 3,743 (32.0%) | 4,200 (30.7%) |  |
| Moderate | 21,984 (48.7%) | 9,566 (48.5%) | 5,725 (49.0%) | 6,693 (48.9%) |  |
| Severe | 8,596 (19.1%) | 3,580 (18.1%) | 2,217 (19.0%) | 2,799 (20.4%) |  |
| Smoking Status |  |  |  |  | <0.001 |
| Current | 13,480 (29.9%) | 6,214 (31.5%) | 3,079 (26.4%) | 4,187 (30.6%) |  |
| Former | 16,482 (36.5%) | 7,081 (35.9%) | 4,484 (38.4%) | 4,917 (35.9%) |  |
| Never | 6,853 (15.2%) | 2,832 (14.4%) | 1,842 (15.8%) | 2,179 (15.9%) |  |
| Unknown | 8,294 (18.4%) | 3,605 (18.3%) | 2,280 (19.5%) | 2,409 (17.6%) |  |
| BMI |  |  |  |  | <0.001 |
| <18.5 | 259 (0.6%) | 134 (0.7%) | 36 (0.3%) | 89 (0.7%) |  |
| 18.5-24.9 | 6,391 (14.2%) | 3,327 (16.9%) | 1,350 (11.6%) | 1,714 (12.5%) |  |
| 25-29.9 | 14,536 (32.2%) | 6,859 (34.8%) | 3,649 (31.2%) | 4,028 (29.4%) |  |
| ≥30 | 23,463 (52.0%) | 9,200 (46.6%) | 6,537 (56.0%) | 7,726 (56.4%) |  |
| Unknown | 460 (1.0%) | 212 (1.1%) | 113 (1.0%) | 135 (1.0%) |  |
| Systolic Blood |  |  |  |  | <0.001 |
| <110 | 4,505 (10.0%) | 1,964 (10.0%) | 1,179 (10.1%) | 1,362 (10.0%) |  |
| 110-119 | 5,855 (13.0%) | 2,589 (13.1%) | 1,478 (12.6%) | 1,788 (13.1%) |  |
| 120-129 | 8,823 (19.6%) | 3,917 (19.9%) | 2,339 (20.0%) | 2,567 (18.7%) |  |
| 130-139 | 11,632 (25.8%) | 5,225 (26.5%) | 3,024 (25.9%) | 3,383 (24.7%) |  |
| 140-149 | 6,335 (14.0%) | 2,744 (13.9%) | 1,603 (13.7%) | 1,988 (14.5%) |  |
| 150-159 | 3,998 (8.9%) | 1,712 (8.7%) | 1,032 (8.8%) | 1,254 (9.2%) |  |
| ≥160 | 3,766 (8.3%) | 1,488 (7.5%) | 976 (8.4%) | 1,302 (9.5%) |  |
| Unknown | 195 (0.4%) | 93 (0.5%) | 54 (0.5%) | 48 (0.4%) |  |
| LDL-c |  |  |  |  | <0.001 |
| <70 | 15,353 (34.0%) | 6,449 (32.7%) | 4,407 (37.7%) | 4,497 (32.8%) |  |
| 70-99 | 13,831 (30.7%) | 6,227 (31.6%) | 3,633 (31.1%) | 3,971 (29.0%) |  |
| 100-129 | 6,835 (15.2%) | 3,072 (15.6%) | 1,534 (13.1%) | 2,229 (16.3%) |  |
| ≥130 | 4,605 (10.2%) | 1,995 (10.1%) | 959 (8.2%) | 1,651 (12.1%) |  |
| Unknown | 4,485 (9.9%) | 1,989 (10.1%) | 1,152 (9.9%) | 1,344 (9.8%) |  |
| DM duration |  |  |  |  | <0.001 |
| 1 year | 3,127 (6.9%) | 2,152 (10.9%) | 407 (3.5%) | 568 (4.1%) |  |
| 2 years | 2,711 (6.0%) | 1,788 (9.1%) | 413 (3.5%) | 510 (3.7%) |  |
| 3 years | 2,468 (5.5%) | 1,398 (7.1%) | 510 (4.4%) | 560 (4.1%) |  |
| 4 years | 3,031 (6.7%) | 1,702 (8.6%) | 620 (5.3%) | 709 (5.2%) |  |
| 5-years | 33,772 (74.9%) | 12,692 (64.3%) | 9,735 (83.3%) | 11,345 (82.9%) |  |
| Co-morbidities |  |  |  |  |  |
| Hypertension | 41,948 (93.0%) | 18,173 (92.1%) | 10,952 (93.7%) | 12,823 (93.7%) | <0.001 |
| Arrhythmia | 14,283 (31.7%) | 6,523 (33.1%) | 3,772 (32.3%) | 3,988 (29.1%) | <0.001 |
| Heart failure | 11,701 (25.9%) | 4,873 (24.7%) | 3,088 (26.4%) | 3,740 (27.3%) | <0.001 |
| Coronary artery disease | 21,813 (48.4%) | 9,317 (47.2%) | 5,807 (49.7%) | 6,689 (48.9%) | <0.001 |
| Valvular heart disease | 4,874 (10.8%) | 2,247 (11.4%) | 1,320 (11.3%) | 1,307 (9.5%) | <0.001 |
| Cerebrovascular disease | 11,021 (24.4%) | 4,881 (24.7%) | 2,865 (24.5%) | 3,275 (23.9%) | 0.23 |
| Chronic Kidney | 13,113 (29.1%) | 5,332 (27.0%) | 3,590 (30.7%) | 4,191 (30.6%) | <0.001 |
| COPD | 15,126 (33.5%) | 7,266 (36.8%) | 3,731 (31.9%) | 4,129 (30.2%) | <0.001 |
| Cancer | 6,729 (14.9%) | 3,368 (17.1%) | 1,716 (14.7%) | 1,645 (12.0%) | <0.001 |
| Dementia | 2,094 (4.6%) | 951 (4.8%) | 513 (4.4%) | 630 (4.6%) | 0.21 |
| Depression | 14,588 (32.3%) | 6,256 (31.7%) | 3,496 (29.9%) | 4,836 (35.3%) | <0.001 |
| Chronic liver | 4,131 (9.2%) | 1,977 (10.0%) | 970 (8.3%) | 1,184 (8.6%) | <0.001 |
| Obesity | 15,409 (34.2%) | 6,034 (30.6%) | 4,184 (35.8%) | 5,191 (37.9%) | <0.001 |
| Anemia | 5,374 (11.9%) | 2,550(12.9%) | 1,425 (12.2%) | 1,399 (10.2%) | <0.001 |
| Weight loss | 2,555 (5.7%) | 1,325 (6.7%) | 517 (4.4%) | 713 (5.2%) | <0.001 |

Overall, 75.5% of study patients were active users of a diabetes medication at baseline, and this proportion was higher in those with worse glycemic control (Table 2). Metformin (45.3%) and insulin (39.8%) were the most prescribed medications, with higher use in patients with HgbA1c >8% compared to ≤7% at baseline. Prescriptions of SGLT2i and GLP-1 at baseline were low. Nearly 90% of study patients were receiving an anti-hypertensive and 75% a statin medication at baseline.

**Table 2.** Baseline Medication Use.

|  | Overall Cohort | Categories of Baseline HgbA1c |  |  |  |
| --- | --- | --- | --- | --- | --- |
|  | N=45,109 | ≤7%<br>N=19,732 | >7% to 8%<br>N=11,685 | >8%<br>N=13,692 | P<br>value |
| **No. of DM medications** | 1.7 (0.8) | 1.3 (0.6) | 1.8 (0.8) | 1.9 (0.9) | < 0.001 |
| Any DM medications | 34,044 (75.5%) | 10,810 (54.8%) | 10,530 (90.1%) | 12,704 (92.8%) | < 0.001 |
| Metformin | 20,420 (45.3%) | 7,321 (37.1%) | 6,252 (53.5%) | 6,847 (50.0%) | < 0.001 |
| Insulin | 17,966 (39.8%) | 2,739 (13.9%) | 5,620 (48.1%) | 9,607 (70.2%) | < 0.001 |
| Sulfonylurea | 11,746 (26.0%) | 3,311 (16.8%) | 4,278 (36.6%) | 4,157 (30.4%) | < 0.001 |
| SGLT-2 | 1,172 (2.6%) | 126 (0.6%) | 392 (3.4%) | 658 (4.8%) | < 0.001 |
| GLP-1 | 1,229 (2.7%) | 140 (0.7%) | 402 (3.4%) | 687 (5.0%) | < 0.001 |
| Others | 3,777 (8.4%) | 747 (3.8%) | 1,376 (11.8%) | 1,654 (12.1%) | < 0.001 |
| **No. of BP medications** | 2.6 (1.3) | 2.6 (1.2) | 2.7 (1.3) | 2.6 (1.3) |  |
| Any BP medications | 40,005 (88.7%) | 17,312 (87.7%) | 10,608 (90.8%) | 12,085 (88.3%) | < 0.001 |
| **Active Statin User** | 33,904 (75.2%) | 14,267 (72.3%) | 9,170 (78.5%) | 10,467 (76.4%) | < 0.001 |
| Low/Moderate Intensity | 17,454 (38.7%) | 7,972 (40.4%) | 4,849 (41.5%) | 4,633 (33.8%) |  |
| High intensity | 16,450 (36.5%) | 6,295 (31.9%) | 4,321 (37.0%) | 5,834 (42.6%) |  |

A total of 8,306 (18.4%) patients experienced a cardiovascular event and 8,199 (18.2%) patients experienced a limb event, with an overall incidence of 4.9 per-100 patient years for both endpoints (Table S3). In multivariable Cox regression analysis with time-varying HgbA1c, cumulative metabolic exposure to hyperglycemia was associated with a significantly increased hazard of clinical events (Figure 1, full model results in Figure S2). Compared with HgbA1c ≤7% (reference), the relative hazard of CV events was 12% higher in patients with HgbA1c >7% to ≤8% and 38% higher in those with HgbA1c >8%. The association between HgbA1c categories and limb events was even stronger, with a relative 20% and 60% higher hazard of events with cumulative exposure to HgbA1c >7% to ≤8% and HgbA1c >8% respectively when compared to HgbA1c ≤7%. This association was consistent when CLTI and major amputation were modeled separately, with a strong relative hazard of major amputation in those with HgbA1c >8% compared to ≤7%. (HR 1.93; 95%CI 1.71-2.18, Table S4)

**Figure 1.**
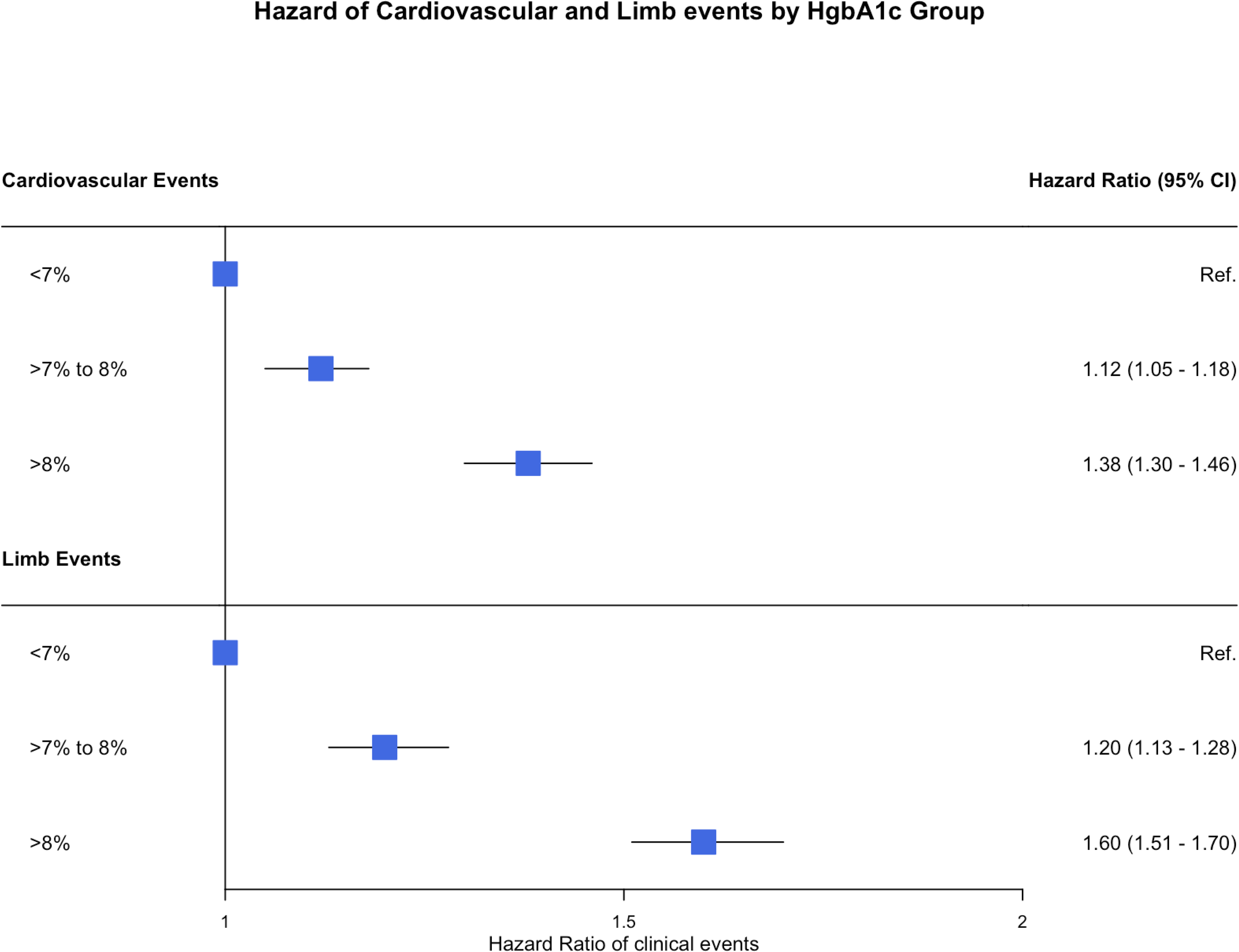
Association Between Cumulative Metabolic Exposure to Hyperglycemia and Risk of Cardiovascular and Limb Events. The forest plot shows risk-adjusted hazard ratios from a multivariable Cox regression model of the association of time-varying hemoglobin A1c and risk of cardiovascular (upper panel) and limb events (lower panel). The model adjusts for age, sex, race, smoking status, PAD severity, duration of diabetes, baseline LDL-C, systolic blood pressure, BMI, baseline medications, and all co-morbidities listed in *Table 1*. The reference HgbA1c category is ≤7%

In absolute terms, the 3-year adjusted incidence of CV events was lowest (11.3%) in those with cumulative exposure to HgbA1c ≤7%, increasing to 12.5% with HgbA1c of >7% to ≤8% and 15.1% with HgbA1c >8%. Similarly, the adjusted 3-year incidence of limb events was 11.1%, 13.2% and 17.0% for cumulative exposure to HgA1c in the ≤7%, >7% to ≤8% and >8% categories, respectively (Figure 2).

**Figure 2.**
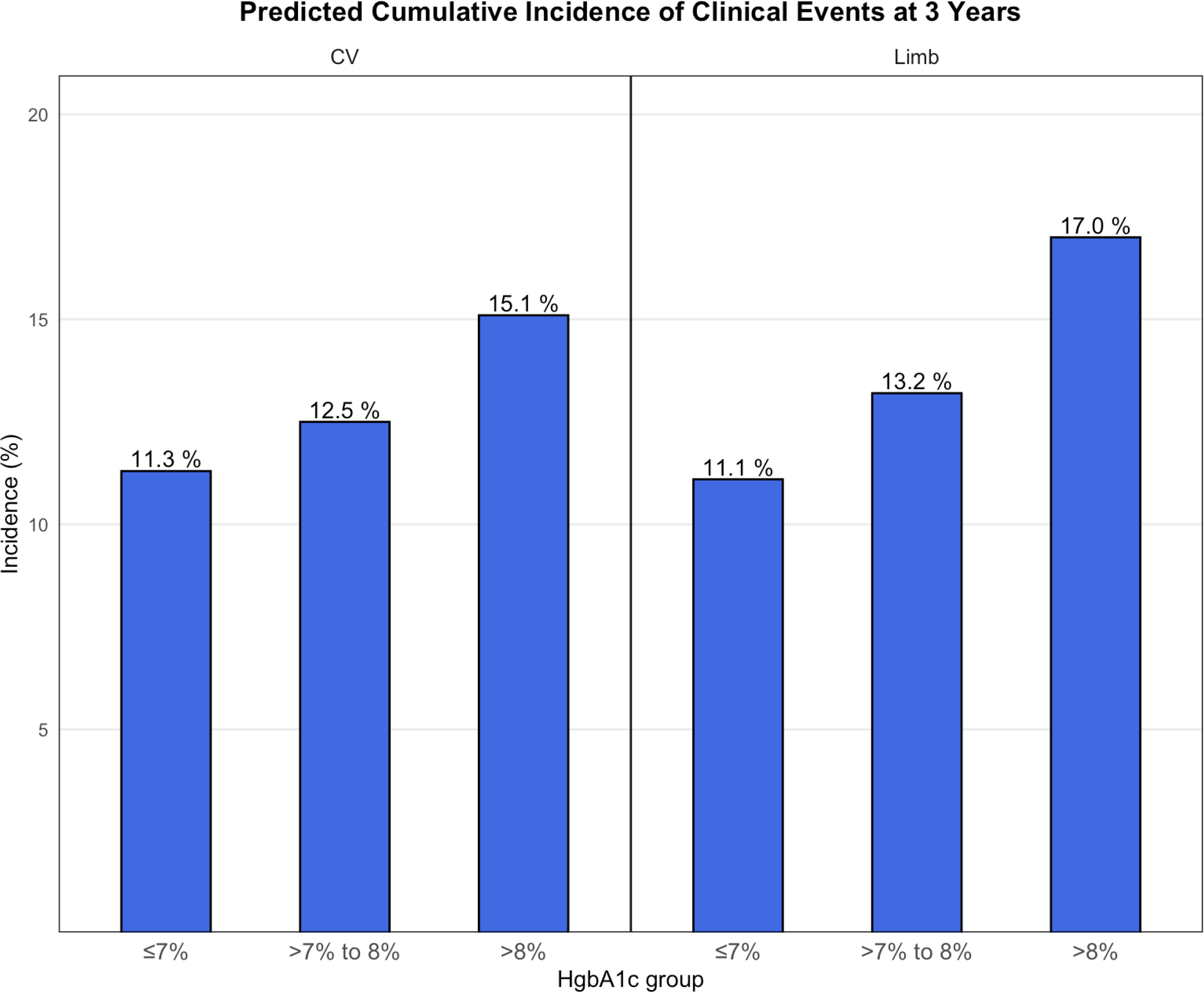
Model Predicted Incidence of Cardiovascular and Limb events at 3 years. We applied the method of recycled predictions to the parameter estimates from the Cox model to quantify the marginal effect of exposure to different HgbA1c categories on the 3-year incidence of cardiovascular (left panel) and limb (right panel) events. For each HgbA1c category, this method fixes the HgbA1c for all patients in the cohort to that category while holding all other variables constant. For example, if the entire cohort experienced a HgbA1c of >8%, the model predicts the incidence of CV events at 15.1% and limb events at 17.0% at 3 years.

In analysis stratified by median age (70 years), we found no significant age interaction between cumulative metabolic burden of hyperglycemia and the risk of CV events and limb events (P for interaction >0.05 for both; Figure 3A). In contrast, when stratifying by severity of PAD, the relative association between HgbA1c categories and limb events was stronger in those with non-severe PAD compared to those with severe PAD (P for interaction <0.01; Figure 3B). However, on an absolute scale, the incremental risk of limb events associated with poor glycemic control was still higher in those with severe PAD, with 3-year adjusted incidence of limb events of 28.8% in HgbA1c >8% vs. 21.3% in HgbA1c ≤7% (absolute difference: 7.5%). In contrast, in those with non-severe PAD, the absolute difference in 3-year cumulative incidence of limb events between HgbA1c >8% vs. Hgb A1c ≤7% was smaller (15.0% vs. 9.2%; absolute difference: 5.8%, Figure 4).

**Figure 3A.**
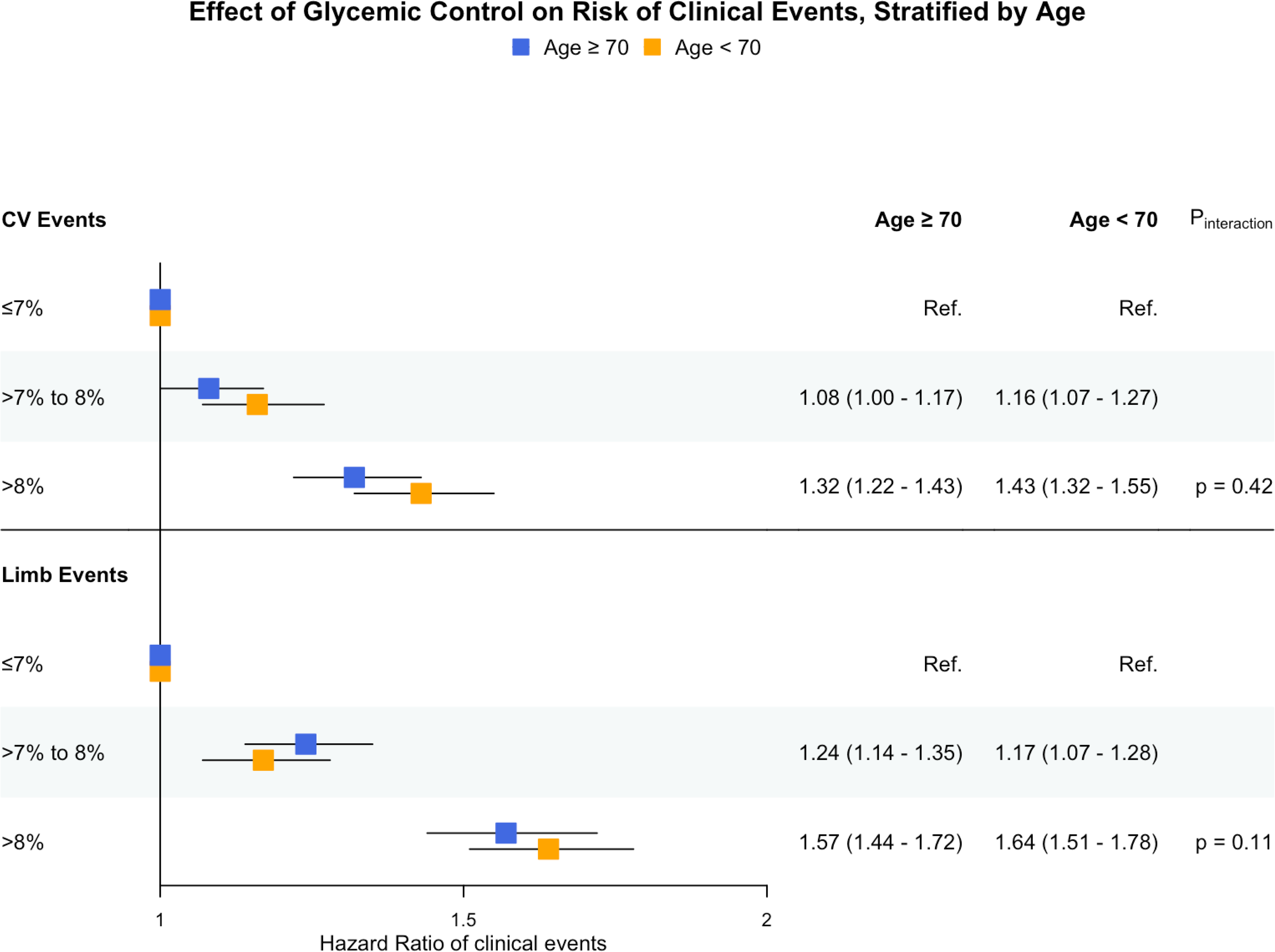
Association Between Cumulative Metabolic Burden of Hyperglycemia with Risk of Cardiovascular (CV) and Limb Events, Stratified by Median Age. The forest plot shows risk-adjusted hazard ratios from separate multivariable Cox regression models of the association between time-varying hemoglobin A1c and the risk of cardiovascular (upper panel) and limb events (lower panel) in patients with age < 70 years (represented in orange) and ≥ 70 years (represented in blue). Both models adjust for age, sex, race, smoking status, PAD severity, duration of diabetes, baseline diabetes medication use, baseline LDL-C, systolic blood pressure, BMI, baseline medications, and all co-morbidities listed in *Table 1*. A threshold of 70 years was chosen as this was the median age. The reference HgbA1c category is ≤7%

**Figure 3B.**
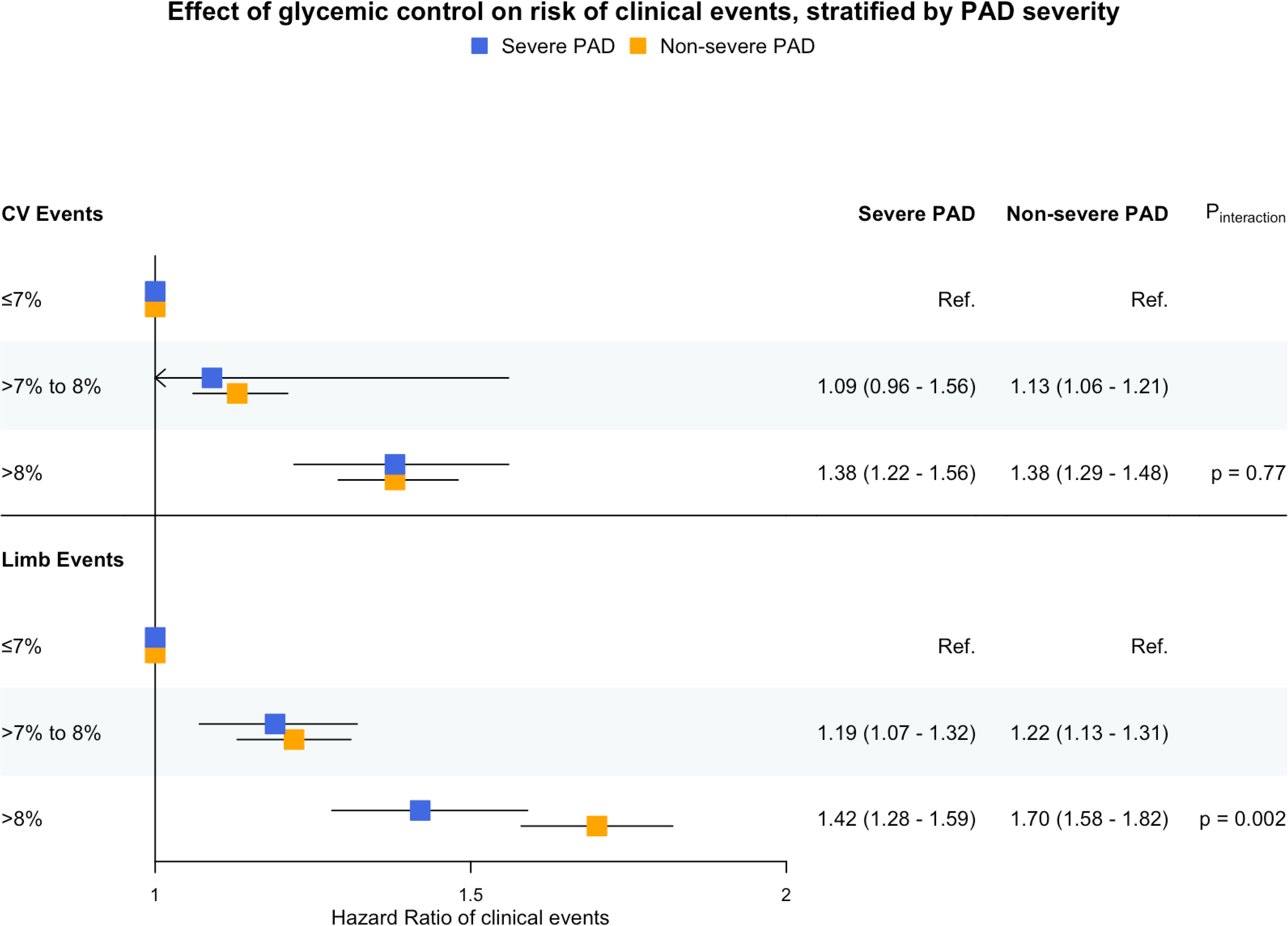
Association Between Cumulative Metabolic Burden of Hyperglycemia with Risk of Cardiovascular (CV) and Limb Events, Stratified by PAD Severity. The forest plot shows risk-adjusted hazard ratios from separate multivariable Cox regression model of the association between time-varying hemoglobin A1c and the risk of cardiovascular (upper panel) and limb events (lower panel) in patients with non-severe PAD (represented in orange) and severe PAD (represented in blue). Both models adjust for age, sex, race, smoking status, PAD severity, duration of diabetes, baseline diabetes medication use, baseline LDL-C, systolic blood pressure, BMI, and all co-morbidities listed in *Table 1*. Severe PAD is defined as 0 ≤ ABI < 0.5 or 0 ≤ TBI < 0.4 in the worst limb. The reference HgbA1c category is ≤7%.

**Figure 4.**
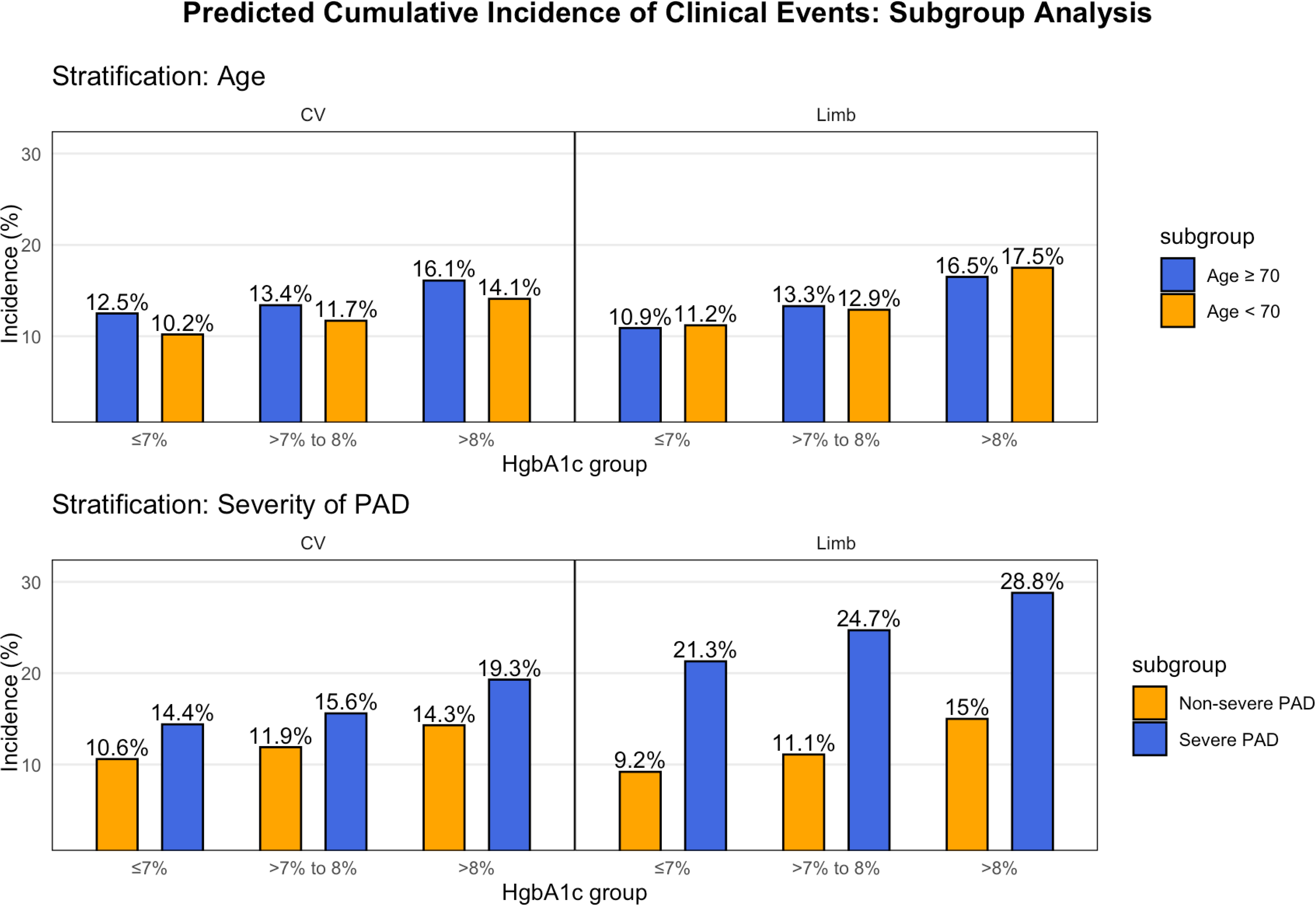
Model Predicted Incidence of Cardiovascular and Limb Events at 3-Years Stratified by Median Age and PAD Severity. Using the method of recycled predictions on the parameter estimates from the Cox model, we quantified the marginal effect of exposure to different HgbA1c categories on the 3-year adjusted incidence of cardiovascular (left panel) and limb (right panel) events, separately for patients age < 70 years vs. ≥ 70 years and non-severe vs. severe PAD. For each HgbA1c category, this method fixes the HgbA1c for all patients in the cohort to that category while holding all other variables constant. Severe PAD is defined as 0 ≤ ABI < 0.5 or 0 ≤ TBI < 0.4 in the worst limb.

In sensitivity analysis, we modeled HgbA1c values as a monthly (rather than 24-month rolling average) time-varying exposure and found that the relative association of time-varying HgbA1c with both CV and limb events was similar (Full model results: Figure S3).

## Discussion

In this study of over 45,000 veterans with new diagnosis of PAD and preexisting diabetes, we found a graded positive association between cumulative metabolic exposure to hyperglycemia and risk of clinical events, with a stronger association observed for limb events compared to CV events. Importantly, the 3-year adjusted incidence of clinical events exceeded 15% in those with cumulative exposure to HgbA1c >8%, compared to 10% in HgbA1c <7%. Given that diabetes is associated with a marked increase in the risk of CV and limb events among patients with PAD,^2,12^ our findings support the need for a clinical trial to investigate the role of intensive glycemic control in these high-risk patients.

Prior RCTs have found a mixed effect of intensive glycemic control on risk of CV events in patients with diabetes. The Action to Control Cardiovascular Risk in Diabetes (ACCORD) trial found no benefit of intensive glycemic control in reducing the risk of cardiovascular events among 10,251 diabetic patients (HR 0.90; 95% CI: 0.78-1.04; P=0.16). In contrast, the United Kingdom Prospective Diabetes Study (UKPDS) and the Veterans Affairs Diabetes Trial (VADT) both demonstrated a modest reduction in cardiovascular events after follow up was extended and while HgbA1c curves remained separated.^7,19^ A meta-analysis that combined data from the above RCTs found an overall 10% reduction in risk of CV events with intensive glycemic control in patients with diabetes, primarily driven by a 15% reduction in risk of myocardial infarction.^8^ However, the overall representation of PAD patients in these clinical trials is low. Moreover, none of the trials adjudicated limb events, a key endpoint in PAD which can be exacerbated by the cumulative metabolic burden of hyperglycemia. Accordingly, there is uncertainty regarding whether the above findings are generalizable to patients with PAD.

Thus, our findings fill an important knowledge gap regarding the effect of cumulative metabolic exposure to hyperglycemia in PAD patients. Compared to HgbA1c ≤7%, exposure to HgbA1c >8% was associated with a marked increase in the risk of cardiovascular and limb events. These findings are consistent with a post-hoc analysis of ACCORD, which found a 30% decrease in risk of amputation in those randomized to the intensive glucose control arm.^20^ Similarly, a post-hoc study using UKPDS data found a statistically significant reduction in the risk of amputation or death from peripheral vascular disease for each 1% reduction in HgbA1c.^21^ However, the limb event rates were low in both studies, due to the under-representation of patients with PAD. In contrast, our study included >45,000 patients with new-onset PAD and diabetes who experienced >8,000 major amputation and CLTI events over a median follow-up of 4.2 years, with an overall incidence of 4.9 limb events per-100 patient-years.

Our results are also consistent with prior studies that have showed increased risk of CV and limb events specifically in those with PAD and diabetes.^9,10,12^ However, these studies have been limited to patients who underwent lower extremity revascularization and only considered HgbA1c at baseline. Given that levels of glycemic control change over time, and the adverse effects of hyperglycemia are cumulative, we build on this prior work by including sequential HgbA1c measures in our analysis. We also noted substantial variability in HgbA1c categories over the duration of follow-up, which underscores the importance of modeling HgA1c as time-varying exposure in risk models. Other notable strengths include the novel approach to PAD identification which used a validated NLP model with high accuracy compared to conventional ICD code approaches,^22–24^ inclusion of rich data on several important confounders, and comprehensive follow-up achieved through linkage of an array of VA and non-VA data sources.

The advent of newer diabetes medications, especially GLP-1 and SLGT2i agents represent a landmark shift in the management of vascular risk in those with diabetes.^25,26^ Multiple recent clinical trials have showed that GLP-1 and SGLT-2 inhibitors reduce the risk of cardiovascular events in patients with diabetes, and emerging data show that GLP-1 agents may reduce limb event risk in patients with established PAD.^27–29^ The benefits of these drugs are believed to be due to a combination of metabolic effects including improved glycemic control.^30^ In our study, metformin was the most commonly prescribed agent, while the use of GLP-1 and SGLT2i at baseline was low, which precludes our ability to evaluate the effect of these drugs. Importantly, we adjusted for diabetes medications in our models, suggesting that the association of poor glycemic control and risk of clinical events could be independent of drug effects.

Clinical practice guidelines permit less stringent HgbA1c goals for older diabetic adults due to the concern of hypoglycemia-related morbidity. Similarly, glycemic control may be de-emphasized in patients with more advanced PAD and competing sources of morbidity.^31^ Despite these considerations, we found that the association between HgbA1c categories and risk of CV and limb events was strong and remained consistent across subgroups stratified by age. For patients with severe PAD, even though the relative hazard of events associated with hyperglycemia may be smaller compared to those with non-severe PAD, the absolute difference in predicted risk of CV and limb events was larger across categories of cumulative exposure to hyperglycemia. This suggests that patients with advanced PAD could still benefit from tight glycemic control. Optimal HgbA1c targets may require an individualized approach, as the burden of clinical events is high in these groups.

Our results should be framed in the context of the following limitations. First, because our study was conducted in the VA, fewer than 3% of participants in our study were women. While the proportion of Black patients was higher than the national average, only 4% of our cohort self-identified as Hispanic. While extrapolation of our findings to these subgroups should be done with caution, there is no reason to believe that the effect of glycemic exposure differs by sex or ethnicity. Second, although we included HgbA1c as a time-varying exposure, demonstrated consistent findings in sensitivity analyses, and adjusted for several confounding variables which included comorbidities, smoking status, severity of PAD, baseline diabetic medication use, blood pressure, BMI, and LDL-cholesterol levels, there is potential for unmeasured confounding. Third, we included participants regardless of whether they were followed within the VA or at outside institutions but only included HgbA1c values available in VA data. These missing data could affect the strength of the demonstrated associations.

In conclusion, we found that longitudinal exposure to poor glycemic control was associated with a markedly increased risk of CV and limb events in patients with diabetes and PAD. Given the high burden of clinical events in these patients, and their relative under-representation in prior landmark clinical trials, our study helps quantify the role of cumulative exposure to hyperglycemia on atherosclerotic vascular disease in this high-risk population of patients with diabetes and PAD.

## Data Availability

Data will be available on a case-by-case basis after discussion with the corresponding author.

